# Longitudinal DNA Methylation Profiling of the Rectal Mucosa Identifies Cell-specific Signatures of Disease Status, Severity and Clinical Outcomes in Ulcerative Colitis

**DOI:** 10.1101/2022.01.28.22269598

**Authors:** Suresh Venkateswaran, Hari K Somineni, Jason D. Matthews, Varun Kilaru, Jeffrey S Hyams, Lee A Denson, Richard Kellamayer, Greg Gibson, David J Cutler, Karen N Conneely, Alicia K Smith, Subra Kugathasan

## Abstract

In peripheral blood, DNA methylation (DNAm) patterns in inflammatory bowel disease patients reflect inflammatory status rather than disease status. Here, we studied DNAm in diseased rectal mucosa from ulcerative colitis (UC) patients, conducting a cell-type-specific EWAS in epithelial, immune and fibroblast cells to understand DNAm changes across disease states, course, and clinical outcomes. At diagnosis, rectal mucosa in UC exhibited a lower proportion of epithelial cells and fibroblasts, and higher proportion of immune cells, along with 3504, 910, and 2279 altered DNAm sites as detected in our cell-specific EWAS, respectively. While treatment had significant effects on DNAm of immune cells, its effects on fibroblasts and epithelial cells were attenuated. Those requiring colectomy exhibited cell composition and DNAm patterns at follow-up more like disease onset than patients who did not require colectomy. Collectively DNAm and gene expression analysis suggest that targeting epithelial genes involved in barrier function may improve clinical outcomes.

## Background

Ulcerative colitis (UC) is a form of inflammatory bowel disease that affects an estimated 1% of the population in North America and Europe^1^. The chronic inflammation is limited to colon in UC, and is usually remitting and relapsing in nature, but repeated inflammation of the colon invariably results in progressive tissue damage. By nature, UC subjects exhibit evidence of systemic inflammation during the active disease, and ∼10% of all UC subjects also have involvement of extra-intestinal manifestations^2^. Although UC is heterogeneous in disease course and outcome, the need for colectomy shortly after diagnosis is the least favorable outcome. Genome-wide association studies have proven the role of immune-associated genetic variants beyond doubt in UC^3-5^, but in total these variants have accounted for less than 10% of the disease susceptibility, and an even smaller proportion of disease outcome. So far, these genetic studies have pointed to few influences of disease beyond the genetic regulation of the immune system itself. Although several drugs are already available to target the immune system in UC, and more immune targeting therapies are being developed, nearly half of UC patients show insufficient response to these therapies. Thus, we asked if it is possible to identify therapeutic targets for UC other than the immune system itself.

Since genetics itself has pointed largely at the immune system, we turned to other molecular systems implicated in UC pathogenesis^6^. DNA methylation (DNAm) signatures^7-9^ have been associated with UC phenotypes. However, to date the majority of DNAm studies in IBD have focused on study of peripheral blood^10-15^. We have shown that DNAm changes in peripheral blood cells associate primarily with inflammatory status rather than disease status^16^, and that methylation patterns in blood largely return to “normal” after anti-inflammatory treatment, regardless of the underlying disease state. To find cells whose molecular signatures might better reflect the disease state, we move to the location of disease itself. Here we examine genome-wide DNAm of the rectal tissue in an inception cohort of UC at 2 time points, once at diagnosis (treatment naïve) and subsequently at follow-up, to explore how longitudinal DNAm associates with disease onset, disease progression and outcome. Similar studies have been done only in small sets of patients, and in select cellular compartments such as purified epithelial cells^17,18^.

Site-specific DNAm differences have been reported in IBD from peripheral blood^16^, and intestinal biopsies^17-19^, but the analysis on the source of the cell type which driving those signals, the temporal relationship between DNAm, disease, and the most unfavorable clinical outcomes (e.g., colectomy status) were not studied for therapeutic benefits. Thus, in this study we conducted a cell type-specific, epigenome-wide association study (EWAS) of DNA methylation (∼850,000 sites) changes in the rectal mucosa at diagnosis, follow-up, and across disease phenotypes and clinical outcome trajectories (colectomy and mucosal healing) using rectal mucosal biopsies collected for the PROTECT pediatric UC inception cohort^20-22^. Our analysis examined interactions between DNAm-based estimates of proportions of three major cell components of intestinal tissues - epithelial, immune cells and fibroblasts (as a measure of mesenchyme) – and DNAm to identify cell-specific differential DNAm patterns and associated gene expression from the same patients to evaluate disease status, disease course, disease severity, and colectomy status as clinical outcomes^23^ with the goal of finding patterns consistent with the cause of disease severity (rather than consequence) to serve as potential targets for molecular therapies.

## Results

### Altered DNAm in the epithelial, immune and fibroblast compartments are associated with UC at diagnosis

We used rectal mucosal biopsies to profile DNAm changes associated with UC. Principal component (PC) analysis of DNAm levels at ∼820K CpG sites in the mucosa showed separate clusters for UC at diagnosis (n=211) and controls (n=85) (**Supplementary Fig. 1**). We observed that PC1 and PC2 explained 20.1% and 8% of the variance in DNAm, respectively. Thus, we performed a traditional EWAS and identified 99,989 DNAm sites associated with UC at diagnosis (FDR < 0.05; **Supplementary Fig. 2**). Identification of this large number of sites may reflect inflammation or unmeasured cellular heterogeneity, but is consistent with the high proportion of variation explained in our PC analysis, and suggests the possibility of a strong but complex DNAm signature of UC. To better understand what drives these large-scale differences in DNAm, we next decomposed our bulk signatures into constituent cell-type proportions (epithelial, immune, and fibroblast cells) via the EpiDISH^24^ algorithms.

**Fig. 1a** shows estimated cellular proportions based on DNAm signatures in UC at diagnosis vs. control samples for three primary cell types (epithelial, immune, and fibroblast cells). The estimated cell proportions from mucosal DNAm profiles shows a decrease in the proportion of epithelial cells (*P* < 2.2e-16) as well as fibroblasts (*P* = 5e-07) and increasing proportions of immune cells (*P* < 2.2e-16), changes that are consistent with damaged and inflamed mucosa^25-27^. Cell-specific EWAS between UC at diagnosis (n=211) and controls (n=85) revealed 3,504 (**Fig. 1b** top panel**; Supplementary Table 1A**), 2,279 (**Fig. 1b** middle panel and **Supplementary Table 2a**) and 910 (**Fig. 1c** bottom panel and **Supplementary Table 3a**) differentially methylated CpG sites in epithelial, immune and fibroblast cells, respectively at FDR < 0.05. Overall, these data suggest the possibility that cell-type-specific changes in DNAm in the rectal mucosa can be leveraged to differentiate diseased tissue from healthy individuals as well as and behavior during disease.

**Fig. 1:**
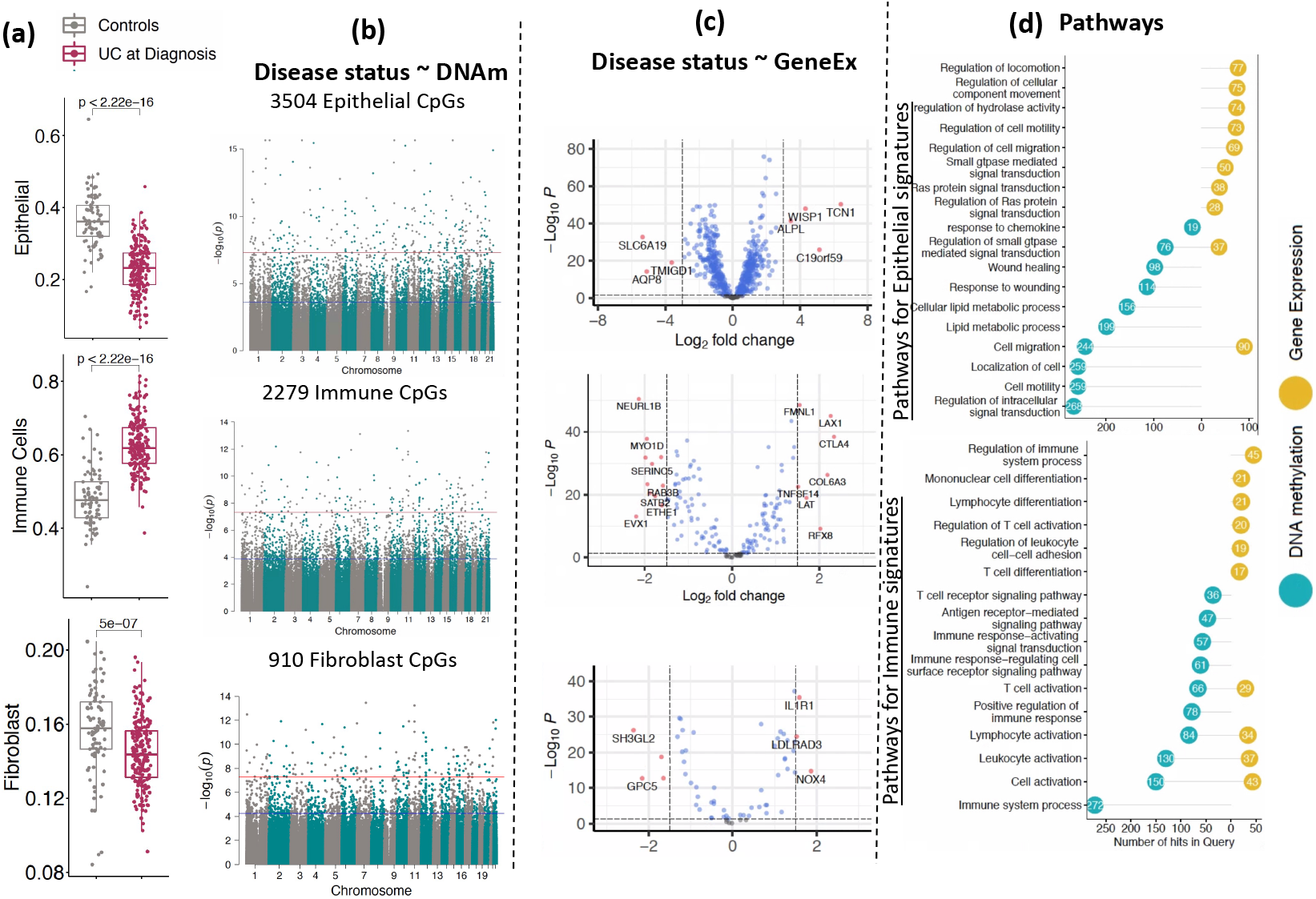
DNAm and corresponding gene signatures associated with UC at diagnosis: (a) Comparison of estimated cell proportions for epithelial, immune cells and fibroblasts between rectal biopsies from UC patients (at diagnosis) and controls. P-values are shown from the Wilcoxon test. (b) Cell-specific epigenome-wide DNAm analysis (EWAS) comparing UC patients to controls. The blue line represents significant differential methylation at FDR < 0.05, and the red line represents Bonferroni-adjusted genome-wide significance (*P <* 1e-08). (c) The volcano plot shows differentially expressed genes that are associated with cell specific CpG sites in UC for all three cell types. x-axis shows log2FC and y-axis shows the negative log p-value detected for each gene in DE analysis. (d) The lollipop diagram shows Gene Ontology (GO) biological processes identified as enriched in sets of differentially methylated CpG sites (blue) and differentially expressed genes in UC. Y-axis shows the number of CpGs and genes detected for each GO term.

### Regulatory potential of the differentially methylated CpG sites in UC

To gain insight into the regulatory potential of our differentially methylated CpG sites, we identified the nearest genes annotated to these CpGs in the Illumina manifest file. Differentially methylated CpG sites in epithelial (n=3504 CpGs), immune (n=2279 CpGs) and fibroblast (n=910 CpGs) cells were annotated to 1932, 1338 and 561 protein-coding genes, respectively. We first examined the association between the differentially methylated CpG sites and the gene expression levels of their corresponding genes in the rectal mucosa (CpG-gene pairs) in a subset of individuals (n=119) with both DNAm and gene expression data, and identified 526 genes associated with 700 CpG sites in epithelial cells (**Supplementary Table 1b**), 173 genes associated with 210 CpG sites in immune cells (**Supplementary Table 2b**) and 63 genes associated with 145 CpG sites in fibroblasts (**Supplementary Table 3b**) at FDR < 0.05. For these sets of genes, we next performed differential expression analysis comparing expression in rectal mucosa biopsies taken from patients with UC at diagnosis (n=206) vs. biopsies taken from non-IBD individuals (n=20). 488 (93%) of the 526 epithelial-specific differentially methylated CpG-associated genes were associated with UC at diagnosis (FDR < 0.05; (**Supplementary Table 1c**). Similarly, 150 (86%) of the 173 immune-specific CpG-associated genes (**Supplementary Table 2c**) and 56 (87%) of the 63 fibroblast-specific CpG-associated genes (**Supplementary Table 3c**) were differentially expressed between patients with UC at diagnosis and non-IBD controls (FDR < 0.05; **Fig. 1c**). The observation that the majority of the genes found to be associated with UC-specific CpG sites also demonstrated changes in steady state levels of mRNA detectable in the mucosa highlights the regulatory potential of our differentially methylated CpG sites.

### Biological relevance of the differentially methylated CpG sites in UC

GO analysis revealed that the set of genes (n=488) associated with the differentially methylated CpG sites in epithelial cells was enriched for numerous biological processes related to epithelial function, including changes in wound response, GTPase signaling and cell migration **(Fig. 1d top panel; Supplementary Tables 1D and 1E)**. Similarly, genes associated with immune cell-specific CpGs (n=150) showed an enrichment for immune function related processes, including cell activation and signaling (**Fig. 1d bottom panel**; **Supplementary Tables 2D and 2E**), all of which are expected to occur in the damaged mucosa ^28-31^. On the other hand, we did not detect any pathway level enrichment for genes associated with fibroblast-specific CpG sites (n=56) at FDR < 0.05.

Taken together, these data suggest that DNAm patterns in different cell populations may be reflecting different aspects of the disease. Immune cell-specific responses are consistent with effects of ongoing inflammation. Immune cell DNAm in rectal tissue, as well as in blood, may be more the consequence of disease caused inflammation than it is the underlying cause of UC or disease progression. The epithelial compartment, on the other hand, with observable differences in wound response and cell migration pathways (prior to treatment) might provide insight into why some individuals fail to respond to effective anti-inflammatory treatments.

### DNAm changes in the rectal mucosa of patients with UC post-treatment

To assess the cellular and molecular changes in the rectal mucosa of patients with UC after treatment, we next examined cellular proportions and DNAm profiles at follow-up (n=73; **Fig. 2**). Clinical activity and/or disease severity in pediatric patients with UC is monitored by a validated activity index known as pediatric UC activity index (PUCAI)^32^. The PUCAI scores of patients at follow-up were significantly lower than those obtained at the time of diagnosis (**Supplementary Fig. 4a)** consistent with patient’s disease activity improving, on average, after treatment. Consistent with this, we observed a corresponding change in the estimated proportions of epithelial, immune and fibroblast compartments **(Fig. 2a)**. Compared to UC at diagnosis, we observe an expansion in the proportion of epithelial and fibroblast compartments at follow-up, and depletion of immune cells, indicating that inflammation has, on average, decreased, and that some mucosal healing was underway.

**Fig. 2:**
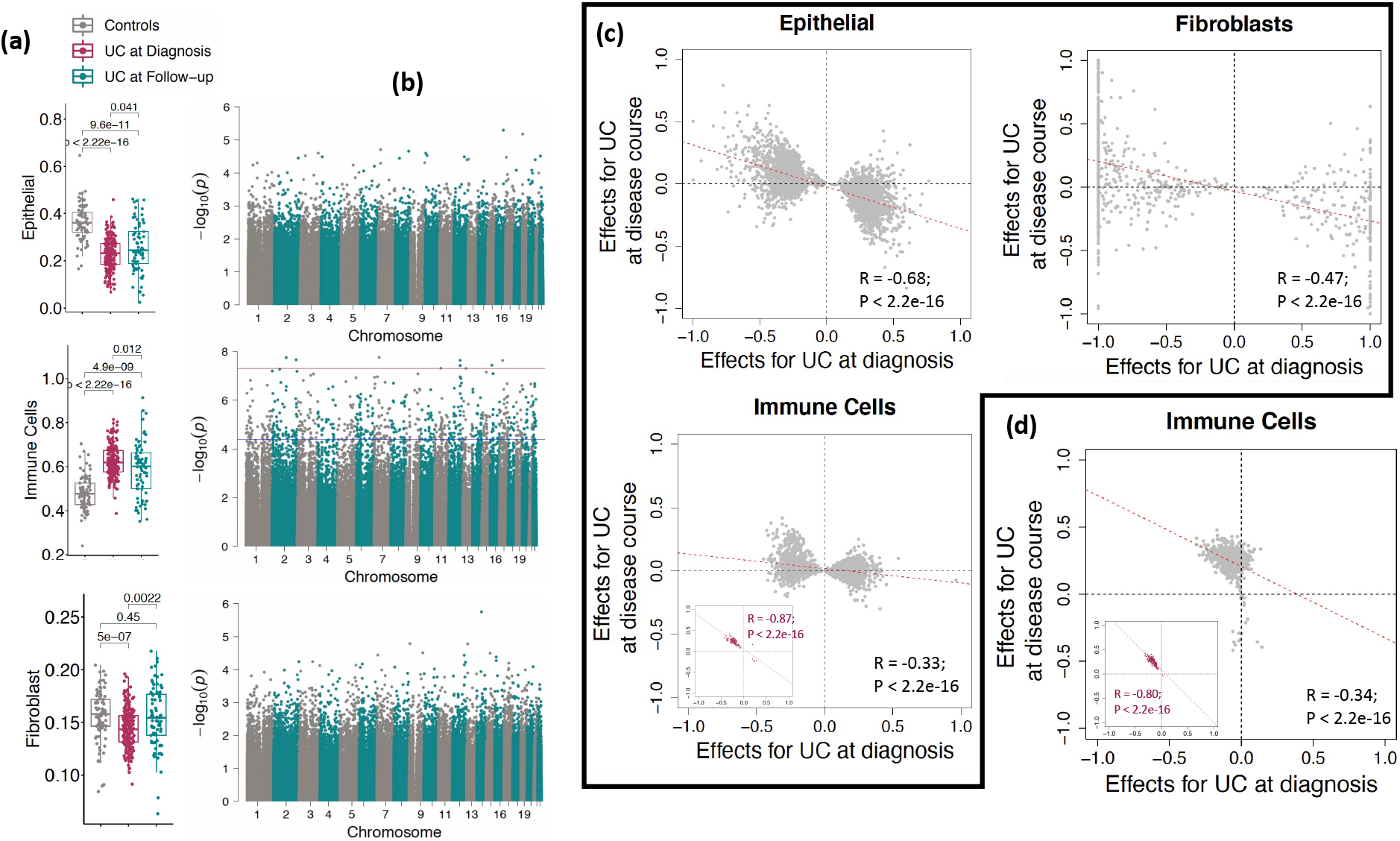
Longitudinal profiling of DNAm in UC patients at during the disease. (a) The boxplots show the rectal biopsy DNAm estimated cell proportions for epithelial, immune and fibroblast compared among controls (n=85), UC at diagnosis (n=211) and UC at Follow-up (n=73). P-values are from the Wilcoxon test. (b) Cell-specific EWAS analysis shows differentially methylated sites between UC at diagnosis and UC at follow-up (paired samples, n=73) for all three major cell types. The blue line represents the sites significant at FDR < 0.05, and the red line represents the sites significant at epigenome-wide *P <* 1e-08. (c) The effect sizes for all the UC associated sites from all three types epithelial (n=3504 CpG sites) and fibroblast (n=910) and immune cells (n=2279) of UC at diagnosis i.e., UC at diagnosis vs Controls (x-axis) were compared to UC at disease course i.e., UC at diagnosis vs follow-up (y-axis). In the immune sub-panel, maroon dots represent the 96 CpG sites that reached significance after multiple test corrections on p-values of 2279 CpGs in UC at disease course (FDR < 0.05). (d) The effect sizes for all the CpG sites associated with disease course i.e., UC at diagnosis vs follow-up (y-axis) from only immune cells (n=668) were compared to UC at diagnosis i.e., UC at diagnosis vs Controls (x-axis). In the sub-panel, maroon dots represent the 154 CpG sites that reached significance after multiple test corrections on p-values of 668 CpGs in UC at diagnosis (FDR < 0.05).

Our cell type-specific EWAS comparing DNAm levels at the time of follow-up to those obtained at the time of diagnosis within the same 73 individuals found 668 CpG sites differentially methylated within the immune compartment (FDR < 0.05; **Fig. 2b** middle panel**; Supplementary Fig. 4c, Supplementary Table 4a**). For the majority of significant sites, the direction of the changes moved DNAm levels in cases after treatment closer to levels seen in controls. These patterns are consistent with DNAm levels in the immune compartment reflecting systemic immune status, and thus responding to systemic anti-inflammatory therapies. On the other hand, EWAS comparing DNAm levels at the time of follow-up to those obtained at the time of diagnosis did not reveal any differentially methylated CpG sites in the epithelial (**Fig. 2b, top panel, Supplementary Fig. 4b**) and fibroblast compartments (**Fig. 2b, last panel, Supplementary Fig. 4d**). This is consistent with epithelial and fibroblast cell populations being less reactive to treatment than the immune compartment.

To identify the impact of treatment effects on DNAm sites, we compared the DNAm differences observed for UC at diagnosis vs. controls, and UC at disease course vs. UC at diagnosis. The EWAS in **Fig. 1b** shows the CpG sites that are differentially methylated between newly diagnosed UC patients and controls, while the EWAS in **Fig. 2d** shows the CpG sites that are differentially methylated in UC patients before and after treatment. **Fig. 2c** shows that for CpGs significant in each of the three compartments in the UC vs. control EWAS (Fig. 1b), there is a negative correlation in effect sizes from the two EWAS; this is consistent with a pattern where CpGs showing DNAm differences in patients at the time of diagnosis return to control levels upon treatment. Stronger negative correlations were observed for sites associated with UC in epithelial cells (R=-0.68) or fibroblasts (R=-0.47) vs. those associated in immune cells (R= -0.33); however, significant pre-vs. post-treatment DNAm differences were only observed in immune cells (**Fig. 2b, middle panel**), with 96 CpG sites showing significant DNAm differences in immune cells in both the case-control and pre-post analyses. For these 96 sites we observed a strong negative correlation (R = -0.86), with DNAm levels of 95 of these sites moving in the opposite direction from onset and trending towards controls and only one site (cg17642041, located in the transcription start site of the *CCR9* gene) trending away from control levels (maroon dot in Fig. 2C immune cell panels). We observed the same general pattern (R = -0.34) at all the 668 CpG sites that differed between diagnosis and follow-up UC: 154 of 668 CpG sites were significantly different at follow-up (FDR < 0.05) with a very strong negative correlation (R = -0.80) between onset and follow-up (**Fig. 2d; Supplementary Table 4e**).

Extending this analysis to all ∼820K CpG sites analyzed across epigenome-wide, we still observed significant negative correlations between effect sizes (R = -0.21 for epithelial cells; R = -0.16 for immune cells; R = -0.21 for fibroblasts; **Supplementary Fig. 5**), suggesting that at a broad scale, the epigenomic level the DNAm profiles observed at follow-up were trending towards those observed in controls. Of the 2279 sites differing between UC at diagnosis and controls in immune cells (from **Fig. 1b, middle panel)**, 33 showed a significant negative association (epigenome-wide) in the comparison of cases vs. controls and a significant positive association when comparing UC at diagnosis vs. follow-up in cases, with a negative correlation between the effect sizes from the two analyses (R = -0.92; **Supplementary Fig. 5b**).

Collectively, these data suggest that between diagnosis and follow-up, the immune compartment undergoes the greatest amount of epigenetic response to treatment, with the epithelial and fibroblast compartments showing fewer changes. These findings are consistent with previous work^17^ reporting that DNAm changes in purified epithelial cells from mucosa persisted across two time-points.

### Biological relevance of the differentially methylated CpG sites in UC at disease course

In support of these patterns showing DNAm profiles at follow-up trending towards control levels, possibly due to inflammation-reducing treatment, gene ontology analysis on the 668 CpG sites differing from UC at diagnosis to follow-up identified numerous biological processes related to immune function, including changes of B-cell/leukocyte activation and differentiation **(Fig. 3a, Supplementary Table 4g)**. To gain further biological insight into the pathways related to DNAm changes in the immune compartment during treatment, we analyzed these 668 CpG sites for association between DNAm and nearby gene expression (270 genes). In this analysis we compared changes in gene expression for each gene with DNAm changes at its associated CpG site across a matched cohort of UC patients (n=29) and observed that expression of 39 genes associated with DNAm at 42 CpG sites (*P <* 0.05; **Supplementary Table 4g**). Differential expression analysis at diagnosis vs. follow-up (n=29 matched samples)^23^ identified 6 of the 39 genes (*FGD2, CRYL1, GXYLT2, PARP1, FOXN3*, and *IL1R2*) as differentially expressed (*P <* 0.05; **Fig. 3b, Supplementary Table 4h**). As an example, we highlight two CpG-gene pairs in **Fig. 3c**, which shows the relative levels of DNAm at CpG sites 1500 base pairs upstream from the transcription start site (TSS) of *FGD2* and *IL1R2* genes, and their corresponding gene expression level for the same UC subjects at diagnosis and follow-up.

**Fig. 3:**
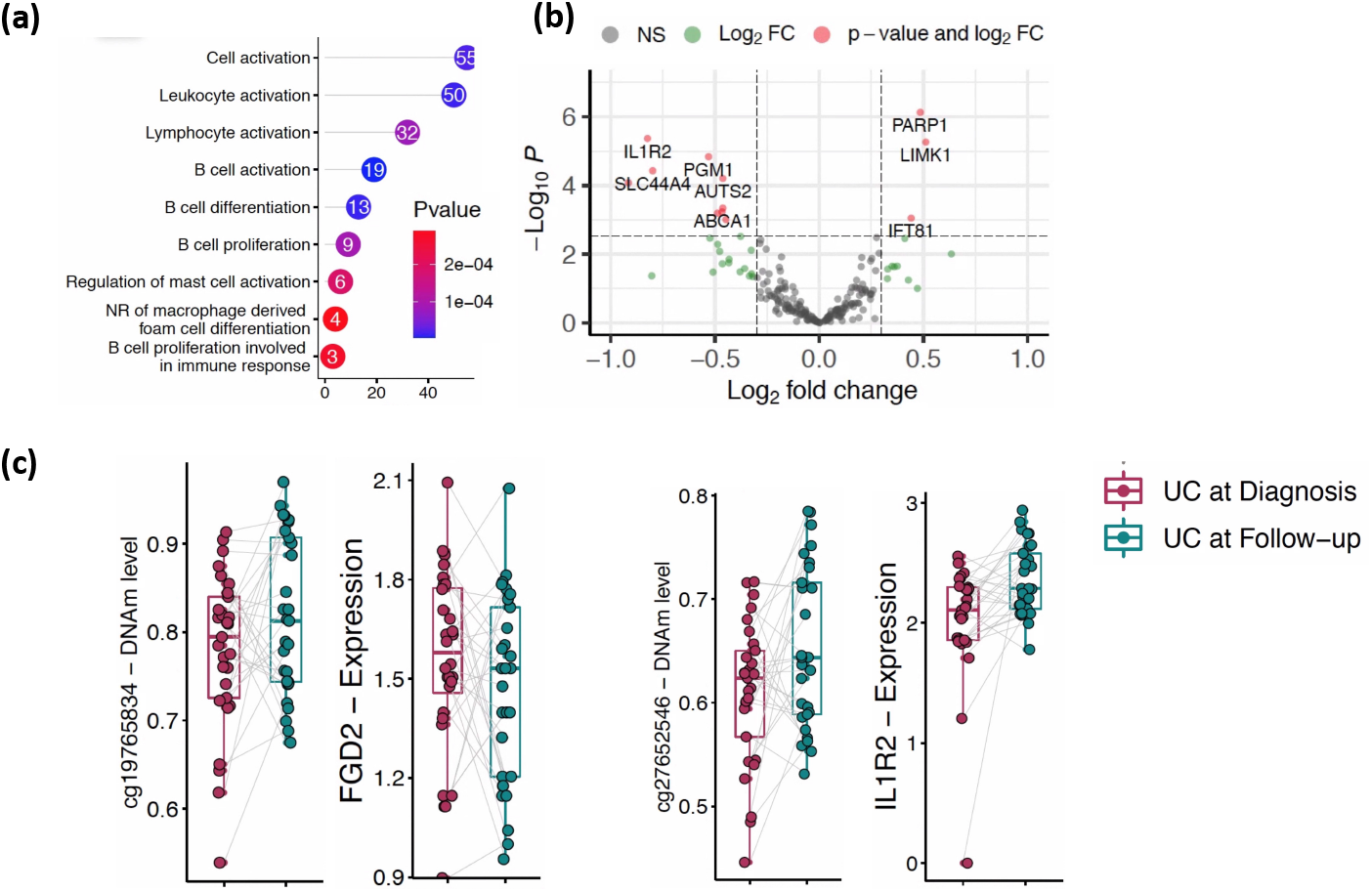
Regulatory importance of the differentially methylated CpG sites in UC at disease course. (a) The lollipop diagram shows Gene Ontology (GO) biological processes identified for 668 differentially methylated CpG sites (blue) from immune cells. Y-axis shows the number of CpGs detected in each GO term. (b) The volcano plot shows differentially expressed genes that are associated with immune cell specific CpG sites in UC during the course of the disease. x-axis shows log2FC and y-axis shows the negative log p-value detected for each gene in DE analysis. (c) Boxplots depicting the methylation proportions of UC at diagnosis and during follow-up (matched sample, n=29, that contains both DNAm and gene expression profiles) at the two CpG sites located TSS1500 to the nearby gene, and the corresponding gene expression values are plotted. Y-axis shows DNAm values and log of read counts representing methylation and gene expression, respectively.

To gain a finer-grained understanding of the immune compartment, we used an immune-subtype reference panel (see methods) to make cell-type composition estimates within this compartment breaking cell-counts into 7 parts (B-, CD8T-, CD4T-cells, monocytes, neutrophils, eosinophils, and nature killer cells). We identified significant changes in the cellular proportions of B-cells and CD4 T-cells between diagnosis and follow-up (*P <* 0.05, Wilcoxon test, **Supplementary Fig. 6**), with decreases in B-cell and increases in T-cell proportions at follow-up. Although neutrophil abundance did not change significantly from diagnosis to follow-up, most of the significant changes in DNAm between diagnosis and follow-up (145 CpG sites) were inferred to be attributable to the neutrophil compartment (**Supplementary Fig. 7; Supplementary Table 4i**). We also observed at least 9 CpG sites showing different DNAm patterns in B-cells (**Supplementary Fig. 7; Supplementary Table 4j)**. Thus, neutrophils, and to some extent B-cells, appear to be undergoing the largest amount of epigenetic change in the mucosa during UC treatment, at least as inferred from this model. Single-cell or purified cell analysis will be the most efficient method moving forward to confirm these model-based inferences.

### UC-specific rectal DNAm signatures at diagnosis correlate with disease severity defined by PUCAI

Next, as a proof-of-concept, we asked if DNAm could distinguish patients with UC based on their disease severity. A comparison of 44 mild UC (10 < PUCAI < 35), 80 moderate UC (35 < PUCAI < 65) and 87 severe UC patients (PUCAI > 65) across the entire 820K CpG panel showed significant differences between mild vs moderate, and mild vs severe UC patients in PC1 **(Supplementary Fig. 8)**. However, no differences were observed between moderate vs severe UC. The estimated cell proportions from mucosal DNAm profiles show no significant differences for either epithelial cell, fibroblasts, and immune cell across disease severity in UC (**Supplementary Fig. 8**). Consistent with this, our cell type specific EWAS among mild, moderate and severe UC patients did not identify any significant CpGs (data not shown). Therefore, to identify individual CpG sites that capture disease severity within each cell type, we grouped patients with moderate and severe UC (n=167) and compared them to those with mild UC (n=44), revealing 46, 127 and 13 CpGs in epithelial, immune and fibroblast compartments, respectively (**Supplementary Fig. 8b, Supplementary Tables 5-7**). GO analysis of these sites did not establish strong evidence for enrichment of any biological processes at FDR < 0.05.

We tested whether any of the cell-specific CpG sites (as a predictor) are associated with nearby gene expression (as an outcome). We observed that only one CpG site (cg10794399) from epithelial cells associated with *AFF3* transcript abundance (**Supplementary Table 5**) and a few CpG sites from immune cells were associated with *C6orf10, PKIB, TUB, KHDRBS3, ACTA2, IRX5, SHANK2, IGFBP6* genes (FDR < 0.05; **Supplementary Table 6**). However, none of these genes were differentially expressed in mild UC when compared to moderate and severe (data not shown).

Collectively, the cell-specific disease severity analysis on the rectal biopsy DNAm from UC at diagnosis indicated that epigenetically moderate and severe UC patients group together, whereas mild UC has a distinct epigenetic profile relative to moderate and severe UC as defined by PUCAI. On the other hand, we couldn’t establish any evidence that these CpG sites are involved in nearby gene regulation or biological processes related to disease severity in UC.

### Rectal DNAm signatures show potential for indicating colectomy risk

The total DNAm signatures were analyzed for signals indicating predisposition towards severe disease resulting in colectomy by year 2 post diagnosis. PC1 of the total 820K CpG sites showed a significant difference in the DNAm signatures between patients who would eventually undergo colectomy at 2 years post diagnosis with patients who did not (**Supplementary Fig. 9**). Similarly, significant differences in DNAm signatures amongst the three compartments, epithelial, immune and fibroblast were also observed between colectomy and no-colectomy. In total, 24 patients underwent colectomy in the 2 years after diagnosis, and while the “no colectomy” group showed improvements in epithelial (*P*= 0.01) and fibroblast (*P* = 0.002) proportions along with a decrease in immune cell proportions at follow-up (*P* = 0.004) (a sign of mucosal healing and reduction of inflammation), the colectomy group showed no improvement in epithelial (*P* = 0.89) and fibroblast proportions (*P* = 0.49) and no decrease in the immune proportions (remaining elevated compared to “no colectomy” at follow-up; *P* = 0.80) (n=175; **Fig. 4a**).

**Fig. 4:**
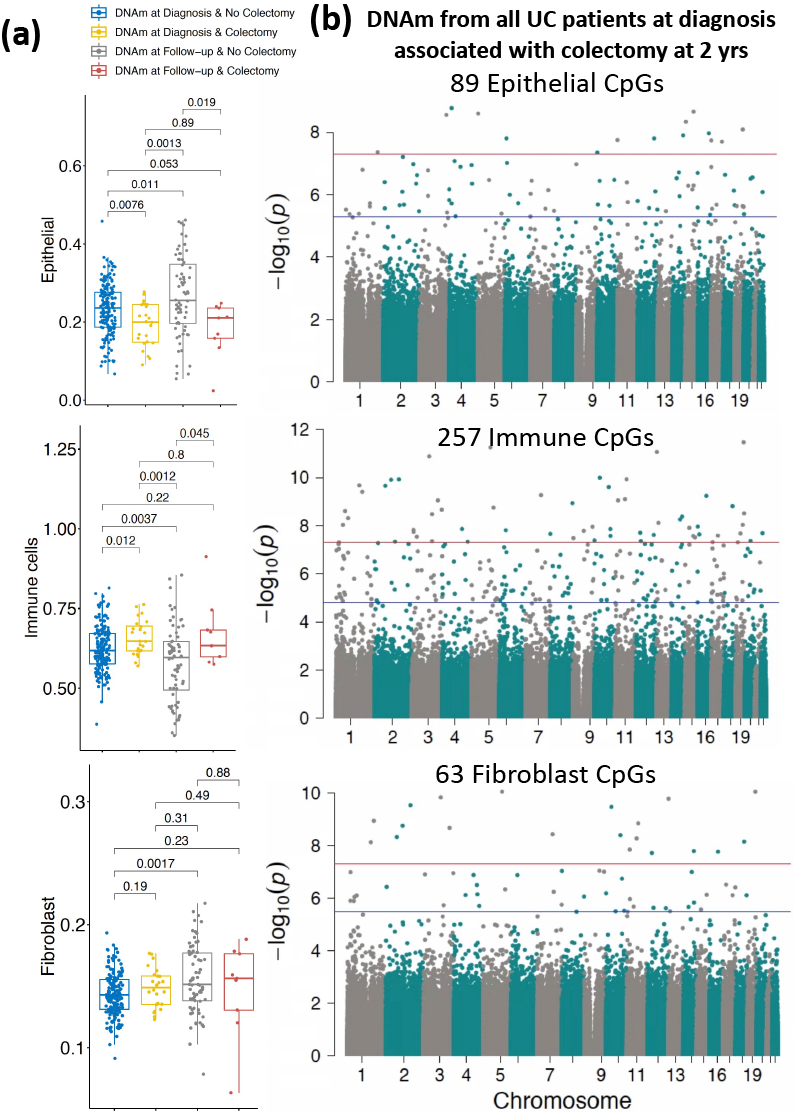
DNAm signatures at diagnosis associated with colectomy at 2 years. (a) Comparison of estimated cell proportions for epithelial, immune cells and fibroblasts obtained from rectal biopsies at diagnosis compared between colectomy and non-colectomy groups. P-values are from the Wilcoxon test. (b) Cell-specific epigenome-wide DNAm analysis (EWAS) comparing UC patients who underwent colectomy at two years (n=24) to non-colectomy UC patients (n=175). The blue line represents significant differential methylation at FDR < 0.05, and the red line represents Bonferroni-adjusted genome-wide significance (*P <* 1e-08).

To identify genes whose DNAm patterns *at diagnosis* indicate the need for future colectomy, cell-type specific EWAS analysis was performed on non-colectomy UC patients (n=175) vs UC patients who eventually underwent colectomy (n=24; **Fig. 4b; Supplementary Fig. 10**). We identified 89 CpG sites from the epithelial cells (**Fig. 4b** top panel**; Supplementary Table 8**), 257 CpG sites from the immune cells (**Fig. 4b** middle panel and **Supplementary Table 9**), and 63 CpG sites coming from the fibroblast (**Fig. 4b** bottom panel and **Supplementary Table 10**) that were different between the colectomy vs. non-colectomy groups. GO analysis of UC colectomy CpG sites did not establish strong evidence for enrichment of any biological processes at FDR < 0.05.

One of the 89 epithelial specific CpGs was associated with *KCND2* expression, but we did not observe any association between expression of *KCND2* and colectomy status. Similarly, 16 out of 257 immune-specific CpG sites were nominally associated with expression of 16 unique genes (**Supplementary Table 9**). Of the 16, only *PFN4* was differentially expressed between colectomy and non-colectomy UC (*P =* 0.04). Lastly, one of the 63 fibroblast-specific CpGs (cg13569868 in *FOXN3*) was associated with significant expression differences, but there was no indication that expression of *FOXN3* is associated with colectomy status in UC. Replication with a considerably larger samples size will be needed for a more convincing demonstration.

We further analyzed differences at diagnosis in the severe UC group by comparing those patients that eventually underwent colectomy at 2 years post diagnosis (n = 14) to those that did not (n = 60). The epithelial cell proportions were nominally decreased *(P* = 0.05) at diagnosis in colectomy severe UC patients when compared to non-colectomy severe UC patients (**Supplementary Fig. 11a, top panel**). In contrast, an increase in the immune and fibroblast proportions was observed for the colectomy group, but the differences did not reach statistical significance (**Supplementary Fig. 11a, middle and bottom panels**). Only 2 CpG sites for epithelial and fibroblast cells were found by cell-specific EWAS analysis (**Supplementary Fig. 11b, top and bottom panels; Supplementary Table 8**,**10**), and 64 CpG sites for immune cells (**Supplementary Fig. 11b**, middle panel**; Supplementary Table 9**). Using a subset of 44 severe UC patients that had both gene expression and DNAm data, we identified 4 of 64 CpG sites from immune cells that were associated with the *PBX3, RHOBTB1, ST3GAL1*, and *ZHX2* genes. Of these, *ZHX2* was differentially expressed between colectomy severe UC and non-colectomy severe UC (p = 0.04). GO analysis of severe UC colectomy associated CpG sites did not establish strong evidence for enrichment of any biological processes at FDR < 0.05.

Collectively, our DNAm estimated cell proportions results suggest that at follow-up, a lack of change in methylation signals with poor indicators at diagnosis (low epithelial, high immune) is a potential clinical indicator for colectomy. Also, joint cell-specific differential methylation and gene expression analysis identified several potential epigenetic sites/genes from immune cells as colectomy-risk biomarkers or possible therapeutic target.

### Clinical relevance of differentially methylated CpG sites in UC

The distinction between methylation signatures in UC at diagnosis and controls suggests that a diagnostic potential exists in these values that can be leveraged for patient stratification. Interestingly, the identified UC specific rectal biopsy CpG sites from epithelial, immune cells, and fibroblasts measured from training dataset, could indeed distinguish UC patients from controls in an independent validation dataset, with 96% accuracy (AUC = 0.96); **Supplementary Fig. 12**). Future studies on DNAm profiled from isolated epithelial, immune cells, and fibroblasts from UC patient’s rectal biopsies are required to establish the diagnostic potential of our cell-specific methylation signatures. On the other hand, we were not able to establish any evidence that the rectal biopsy DNAm profiles predict disease progression or outcome in UC. An experimental design more directed at contrasting UC patients who undergo colectomy at follow-up from non-colectomy UC patients would have more power to find such a signature.

## Discussion

In this study, we used a treatment naïve inception UC cohort in which biospecimens underwent multiomic analysis at baseline and follow-up. Taking advantage of this inception cohort with extensive prospective clinical meta-data, we estimated DNAm-based cell type proportions in the human rectal mucosa – epithelial, immune and fibroblast compartments - and examined DNAm differences in each of these compartments at the time of diagnosis and during the course of UC.

We first showed that within each compartment, UC-specific epigenetic changes are disturbed at diagnosis. We next found that after treatment, overall cell proportions, and many of the cell-specific methylation patterns moved closer to levels seen in controls. This pattern was most pronounced in immune cells and is hardly surprising since current treatment primarily targets the immune system and clearly its effects are most direct and observable in this compartment. Further analysis showed significant differences in estimated cell proportions at follow-up between individuals with mild UC and the moderate/severe group. There were no experiment-wide significant differences in DNA methylation level between moderate and severe disease. On the other hand, there were widespread methylation differences at follow-up between those with ongoing, severe disease who would soon need colectomy, and those who would not. Remarkably, DNAm signatures observed in the rectal mucosa at diagnosis could also distinguish patients destined to need a colectomy within 2 years and those who did not. However, given that differential expression for *PFN4* was only nominally significant, replication attempts of these patterns in larger samples are warranted.

The epithelial barrier is the front-line defense against invading microbes, toxins, and other luminal contents, but also provides selective transport of nutrients and other beneficial substances that maintain homeostasis and carries significant inflammatory consequences to underlying mucosa upon its malfunction and degradation. The lack of epigenetic changes in the epithelial and fibroblast cells following treatment is of concern to mucosal healing. Notably, we observed that at least 25% of the epithelial derived CpG sites (889 out of 3504) changing during UC are consistent with a previously published study that compared purified epithelial cell methylation patterns between UC patients and controls^17^. Damage and erosion of the intestinal epithelium is a hallmark characteristic of UC, and the large number of epithelial-specific CpG sites affected at diagnosis indicates either a response to external stimuli potentiating the disease or a response to the damage that the body is trying to repair. In fact, in the study of epithelial-derived organoids described in the Introduction, at least 25% of the genes (124 out of 488) associated with epithelial derived CpG sites were also observed to be differentially expressed in UC^17^, supporting a relationship between changing DNAm and gene transcription in the epithelium during UC. Our observation of decreased epithelial proportions based on changes in DNAm signals, with a corresponding increase in immune cell abundance, is in line with the well-documented biological consequences of UC. Pathway analysis of the genes influenced by the significantly changing CpG sites showed changes in pathways regulating cell migration, lipid metabolism, small GTPase signaling and wound repair/healing, suggesting that the changes in DNAm in turn influence nearby genes and processes that are critically involved in restitution and repair^33-36^.

Fibroblasts originating from the mesenchymal compartment are responsible for maintaining the extracellular milieu that provides structural stability and regulatory growth and differentiation signals for the epithelium^37-39^. The decrease in proportion of this cellular compartment further correlates with symptoms of UC, but this was the one compartment that looked most similar in comparison to control abundance at follow-up. The epigenetic findings here provide new targets that should be considered for correcting UC related epigenetic changes that could promote mucosal healing. For example, *UBE2G1* detected in fibroblasts was found to be hypomethylated and did not revert after treatment, and is a known IBD marker ^40^. *UBE2G1* is ubiquitin conjugating enzyme involved in degradation of short-lived proteins and known to have disease implications when its functionality is perturbed ^41-43^. Further, whether therapeutic changes in the fibroblasts enhanced their immunosuppressive effects and facilitated a decrease in immune cell proportion was not entirely clear, but a reversion of many of the fibroblasts’ CpG signatures to non-IBD status suggested this possibility.

Clearance of the immune response by current IBD treatments (anti-TNF as an example) has long been appreciated as the standard of care for IBD and was exemplified in our DNAm analysis where healing was associated with a decrease in mucosal immune proportions and reversion of many of the immune-specific CpG sites back to non-IBD signatures. Interestingly, *FOXP1* ^44^, *IL23R*^45-48^, *CCR9*^49,50^ have been documented to play a role in IBD, and here their corresponding CpG loci were found to maintain disease-specific DNAm signatures after treatment. The *CCR9/CCL25* signaling axis is responsible for leukocyte trafficking to the gut and our data may reflect epigenetic changes during UC that are associated with the activation and recruitment of immune cells from the peripheral blood into the intestinal mucosa, consequentially sustaining inflammatory conditions and prolonging or worsening the disease. New targeted therapies that can offset epigenetic effects regulating *CCR9* may show improved resolution of the inflammatory response ^50^.

Additionally, our study has revealed some important cell- and disease-severity specific findings that were not reported in previous literature. In our previous study, we reported that peripheral blood DNAm signatures in IBD associate with the degree of inflammation, but do not predict/characterize disease in the gut. In the current study, utilizing samples provided by the longitudinal UC patients from PROTECT cohort, we were able to address this question, demonstrating that the DNAm patterns from actual diseased tissues reflect the nature of disease rather than the inflammation status. This is further supported by our comparative analysis of cell-specific EWAS indicating that only 9 sites from both epithelium and immune cells, and 4 sites from fibroblasts, overlap with our previous total peripheral blood DNAm sites^16^. Unlike blood DNAm, the rectal tissue DNAm, particularly epithelial / mesenchymal tissue DNAm, do not revert back to normal levels. While contrasts between mild and moderate UC were largely difficult to interpret, differences between individuals requiring early colectomy and those with less severe disease were more pronounced. Methylation patterns at diagnosis seem to predict which patients are most likely to progress to colectomy after two years post treatment, and methylation and cell-composition patterns which fail to revert towards controls at follow-up are especially good predictors of who will require an early colectomy.

Our study is not without limitations. Our analysis was based on correlating clinical parameters with only one disease site (rectum), and the estimated cell proportions and identity are very much model-based estimates that should be confirmed with more direct experimentation approaches as performed in the previous single-cell or purified cell studies ^9, 17,51,52^. The model we employed to decompose mucosal data (comprised of numerous cell types) is believed to be well powered to distinguish three main cellular lineages (epithelial, immune and fibroblasts), but both imprecision in the model estimates as well as loss of sub-type resolution is unavoidable. While our study may be the most cutting-edge analysis that could be done on these tissues when collected at this sample size, we expect large scale-single cell analysis will confirm or reject our inferences in the foreseeable future.

In summary, we show that cell type-specific epigenetic changes are taking place in the rectal mucosa during the course of UC, and these changes are associated with disease severity and outcome. Further, our data also strongly suggest that, in IBD, we should focus studying the relevant tissue for DNAm and for other molecular signatures rather than blood. Based on our findings, we speculate that individuals who do not sufficiently respond to current therapies, targeting epithelial genes with barrier function, perhaps by attempting to revert methylation patterns to control levels, may improve mucosal healing.

## Methods

### Overall study participants

The cases used for analysis here are a subset of the PROTECT UC cohort. PROTECT is a multicenter inception cohort with a total of 431 treatment naïve UC patients from 29 centers in the USA and Canada. This cohort was prospectively followed for at least 2 years with rectal biopsy collection at diagnosis (before treatment) and subsequent follow-up (treatment period from 8 weeks to 2 years; follow-up; n=73). Details regarding inclusion / exclusion criteria, study protocol, approvals, and other clinical parameters assessed have been reported previously^21,22^. Based on the availability of the rectal tissue DNA, 211 pediatric UC patients from the PROTECT cohort, aged 4–17 years old, were used. Out of the 211 had baseline rectal DNA availability, 73 of them also had follow-up rectal tissue DNA and were used. All 73 patient follow-up samples were collected as clinically indicated during the follow-up visits between 8 weeks to 2 years. All 73 patients received one or more treatment(s) prior to follow-up, described in detail in Hyams et al^21,22^. The UC diagnosis was based on conventional clinical, endoscopic, and histological parameters. For non-IBD controls, we used age- and gender-matched rectal biopsy genomic DNA samples from 85 RISK participants (RISK is described elsewhere^20^) that had no histologic or endoscopic inflammation and remained asymptomatic during the disease course. Both the PROTECT and RISK studies were approved by the Institutional Review Boards at each of the participating RISK and PROTECT sites. The same sites in North America participated in both the PROTECT and RISK studies. All relevant ethical regulations for work on human participants have been met and conducted in accordance with the criteria set by the Declaration of Helsinki. Informed consent was obtained from the parents of all study participants.

### Quantification of genome-wide DNAm

Rectal biopsy genomic DNA was extracted using the AllPrep DNA/RNA Mini Kit (Qiagen, Valencia, CA), and 500ng of DNA was subjected to bisulfite treatment using EZ DNAm-GoldTM Kits (Zymo Research, Irvine, CA). MethylationEPIC BeadChip (Illumina, San Diego, CA) was used to quantify genome-wide DNAm differences across ∼850,000 genome-wide CpG sites ^53^. The R package CpGassoc^54^ was used to perform the initial quality control (QC). CpG sites called with low signal or low confidence (detection *P* > 0.05) or with data missing for greater than 10% of samples were removed, and samples with data missing or called with low confidence for greater than 10% of CpG sites were removed. Probes mapping to multiple locations were also removed^55^. After QC, a total of ∼820,000 probes and 369 samples (85 non-IBD controls, 211 UC samples at diagnosis, and 73 samples from follow-up) remained. Beta values (*β*) were calculated for each CpG site as the ratio of methylated (M) to the sum of methylated and unmethylated (U) signals: *β*=M/(M+U). Signal intensities were then normalized using the module beta-mixture quantile dilation (BMIQ)^56^ to account for the probe type bias. These normalized signal intensities were used to perform principal component analysis to further identify sample outliers.

### Estimating the cell proportions of rectal biopsy tissues

The R/Bioconductor-package EpiDISH^24^ was used to estimate cell type proportions through deconvolution of our bulk DNAm data derived from rectal tissue. Estimation was based on two reference panels: (i) an epithelial, immune and fibroblast -specific reference panel and (ii) another immune subtype cells reference panel that can also map the data to 7 immune cell types; B-cells, CD4+ T-cells, CD8+ T-cells, NK-cells, monocytes, neutrophils, and eosinophils. These estimated cell type proportions were used as covariates in all DNAm analyses to adjust for differences in DNAm due to between-sample differences in cellular composition.

### Genotyping and data processing

Peripheral blood DNA samples of 296 cases and controls with DNAm data were genotyped using the UK Biobank array, and ∼850,000 genotypes were called using the Axiom Suite software. All subjects had call rates >95% and consistent gender records with the clinical data. All quality control procedures were performed in PLINK^57^. Principal components were computed based on a pruned version *“--hwe 0*.*001 --maf 0*.*2 --geno 0*.*01 --indep-pairwise 50 5 0*.*2*” of the data set consisting of 58,237 LD-independent SNPs (r^2^ < 0.1). We used the first 5 genotype-based principal components to control for population stratification in all analyses.

### DNAm association with UC at diagnosis

To identify CpG sites associated with UC at diagnosis, we performed a case-control EWAS on 211 UC patients at diagnosis compared to 85 non-IBD controls. UC-associated methylation changes in rectal tissue regardless of cell type were first profiled using the R package CpGassoc^54^. Briefly, DNAm was regressed on disease status (0 for control, 1 for UC) with age, gender, epithelial and fibroblast cell proportions, and the first five genotype-based principal components as covariates in the model.

### Cell-specific methylation association with UC at diagnosis

We next used the CellDMC function from the EpiDISH^24^ package to identify sets of CpGs that show a cell-type specific association with UC. We performed cell-specific EWAS within the epithelial, immune and fibroblasts to test for association between UC and methylation at the ∼820,000 sites that passed QC. DNAm at each CpG was regressed on disease status (0 for control, 1 for UC) and age, gender, and the first five genotype-based principal components were included as covariates in the model, along with covariates for DNAm-based estimates of cellular proportions, and an interaction term between UC and cellular proportion to identify cell-specific signal. We identified significant CpGs within each EWAS using the Benjamini-Hochberg false discovery rate criterion (FDR<0.05).

### Cell-specific methylation association with UC at diagnosis versus follow-up

To assess longitudinal changes in DNAm in patients with UC, we compared the methylation levels in rectal biopsy samples obtained at diagnosis and follow-up (n=73; from 8 weeks to 2 years). Age was recalculated for the follow-up samples based upon their time of visit after diagnosis. In the cell-specific EWAS, DNAm was regressed on disease course (0 for at diagnosis, 1 for at follow-up), and age, gender, and the first five genotype-based principal components were included as covariates in the model, along with the covariates and interactions for cell type proportion. To account for the two time points from each patient, representing DNAm levels at diagnosis and follow-up, we included fixed effect covariates for subject ID. To identify CpG sites associated with UC during treatment, we performed a similar cell-specific EWAS comparing diagnosis vs follow-up within the same samples (n=73).

### Gene Ontology (GO) biological process enrichment analysis

Gene ontology (GO) for biological process enrichment analysis was performed for genes annotated to our UC associated CpG sites by the R/Bioconductor package missMethyl^58^. Genes with more CpG probes on the MethylationEPIC array are more likely to have differentially methylated CpGs, which could introduce potential bias when performing pathway enrichment analysis. The *gometh* function implemented in missMethyl considers the varying number of differentially methylated CpGs by computing a prior probability for each gene based on the gene length and the number of CpGs probed per gene on the array. Similarly, the GO biological process enrichment analysis for UC associated genes that are associated to UC associated CpGs was performed by using Toppgene^59^.

### Analysis of DNAm and gene expression

Differential gene expression analysis was performed on a subset of patients from whom both gene expression and DNAm data was available from the rectal mucosa (n=119). Gene expression was previously measured using TruSeq Illumina mRNAseq (20 controls and 211 UC samples)^23^ or Lexogen 3’UTR mRNAseq (39 UC at diagnosis and a matched subset of the same 39 UC patients’ sampled at 52 week follow-up)^60^. More details on RNA sequencing, data processing and QC are described elsewhere^23,60^.

CpG sites associated with disease status, disease course, disease severity, or colectomy status in the EWAS were further tested for association with expression of genes as annotated by Illumina. In total we used data from 119 patients with UC who had both DNAm and TruSeq Illumina mRNAseq gene expression profiles^23^. DNAm values were regressed on gene expression counts using the *lm*() R function, with age, gender and the first five genotype-based principal components included as covariates in the model. A false discovery rate criterion (Benjamini-Hochberg FDR<.05) was used to define a set of significant CpG-gene pairs.

### Differential gene expression analysis on disease status, disease severity, and colectomy status

We also performed a targeted differential expression analysis of genes annotated to the CpG sites identified in the EWAS of disease status, disease course, disease severity and/or colectomy status. The TruSeq Illumina mRNAseq dataset^23^ (n=119) was used to identify genes differentially expressed between disease status, disease severity and/or colectomy status. Similarly, the Lexogen 3’UTR mRNAseq data was analyzed to identify genes differentially expressed for CpGs associated to UC at disease course. The DEseq2 package was used to perform the differential expression analysis. Briefly, gene expression values were regressed on disease status, disease severity, or colectomy status with age and gender adjusted as covariates in the model using the default normalization method. DE genes were identified using either nominal p-value < 0.05 and fold change (FC) > 1.2.

### Random forest classification

The entire dataset at diagnosis (n=296) was divided into a 75% training (64 controls and 158 UC cases) and a 25% validation set (21 controls and 53 UC cases). The CellDMC was performed only on training dataset samples and the UC specific CpG sites across all three cell-types were identified. A RandomForest (RF) model constructed using the identified UC specific CpG sites of training dataset using randomForest package^61^ in R with default parameters. The trained RF model was tested using the test dataset samples to test the prediction performance of the defined model. Accuracies (ACC) and area under the curve (AUC) were calculated by comparing of actual labels to predicted labels of test set class, separately for cases and controls.

## Data Availability

The DNAm data for all the 369 rectal biopsy samples included in this study have been deposited in the Gene Expression Omnibus (GEO) and are accessible through GEO series accession GSE18506.
To review GEO accession GSE185061:
Go to https://nam11.safelinks.protection.outlook.com/?url=www.ncbi.nlm.nih.gov%2Fgeo%2Fquery%2Facc.cgi%3Facc%3DGSE185061&data=04%7C01%7Csuresh.venkateswaran%40emory.edu%7C608f82f0576743c3699708d9dc2a5235%7Ce004fb9cb0a4424fbcd0322606d5df38%7C0%7C0%7C637782896589960813%7CUnknown%7CTWFpbGZsb3d8eyJWIjoiMC4wLjAwMDAiLCJQIjoiV2luMzIiLCJBTiI6Ik1haWwiLCJXVCI6Mn0%3D%7C3000&sdata=q8AL8lh3%2BJoLySJkL%2FaahHLThr3lil6NXNfSxMwxaAo%3D&reserved=0
Enter token abylkamahxcjlwp into the box

## Abbreviations

(ACC): Accuracies
(AUC): Area Under the Curve
(DNAm): DNA Methylation
(EWAS): Epigenome-Wide Association Studies
(FDR): False Discovery Rate
(GO): Gene Ontology
(IBD): Inflammatory Bowel Disease
(PUCAI): Pediatric Ulcerative Colitis Activity Index
(PROTECT): Predicting Response to Standardized Pediatric Colitis Therapy
(PCs): Principle Components
(RISK): Risk Stratification and Identification of Immunogenetic and Microbial Markers of Rapid Disease Progression in Children with Crohn’s Disease
(UC): Ulcerative Colitis

## Data availability

The DNAm data for all the 369 rectal biopsy samples included in this study have been deposited in the Gene Expression Omnibus (GEO) and are accessible through GEO series accession GSE185061. Metadata details regarding subset of patients used in this study are summarized in **Supplementary Table 11**.

## Acknowledgements

We are grateful to Anne Dodd, and Jarod Prince for their support and helpful comments.

## Author Contributions

K.N.C, A.K.S, H.K.S and S.K. conceived and designed the study. S.V. performed the analysis with input from D.J.C., K.N.C., A.K.S. and G.G. processed samples for methylation profiling. J.S.H., L.A.D. and S.K. participated in the conception and design of the RISK and PROTECT studies. S.V., J.D.M., H.K.S., R. K., G.G., D.J.C., K.N.C., A.K.S., and S.K. interpreted the results and wrote the manuscript. All authors reviewed and approved the manuscript prior to submission.

