## Supplementary Figures for "Longitudinal DNA Methylation Profiling of the Rectal Mucosa Identifies Cell-specific Signatures of Disease Status, Severity and Clinical Outcomes in Ulcerative Colitis"

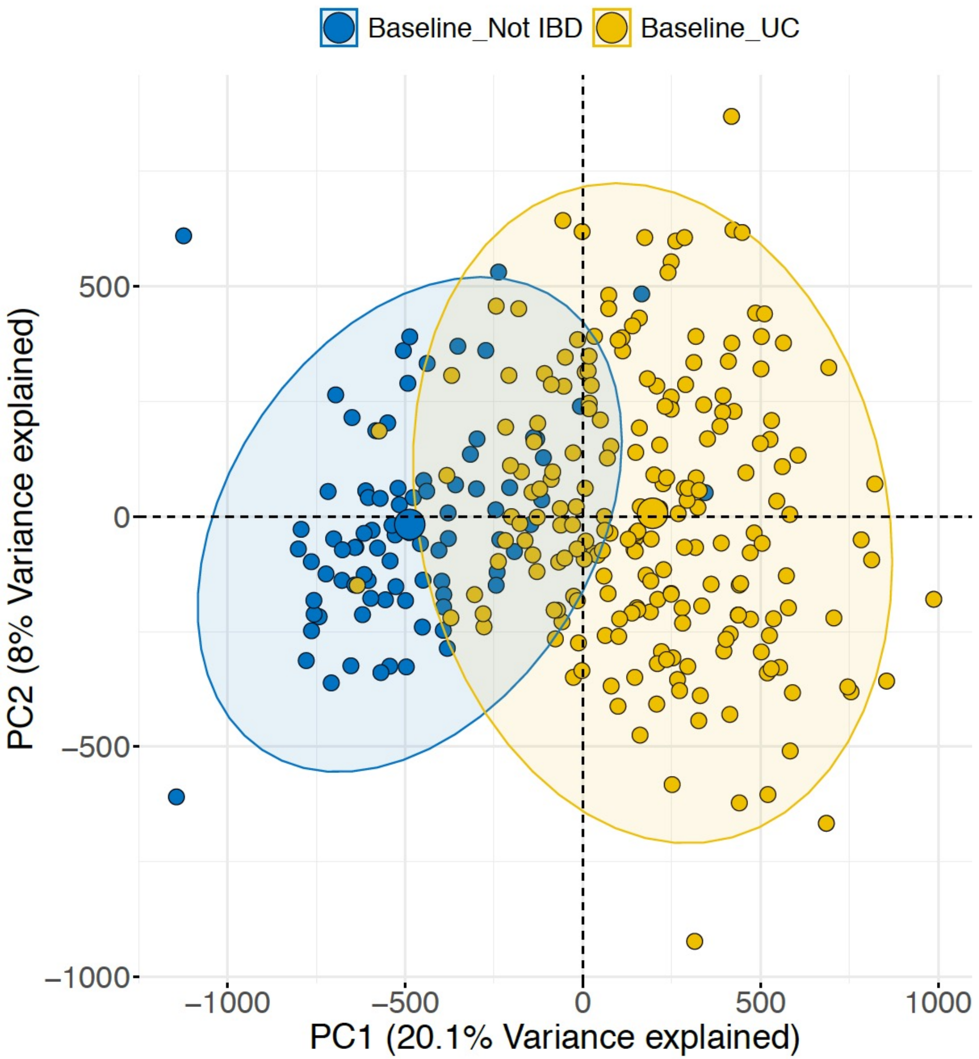


**Supplementary Figure 1: The principal component analysis on entire ~820k sites shows two distinct clusters for controls and UC at diagnosis.**

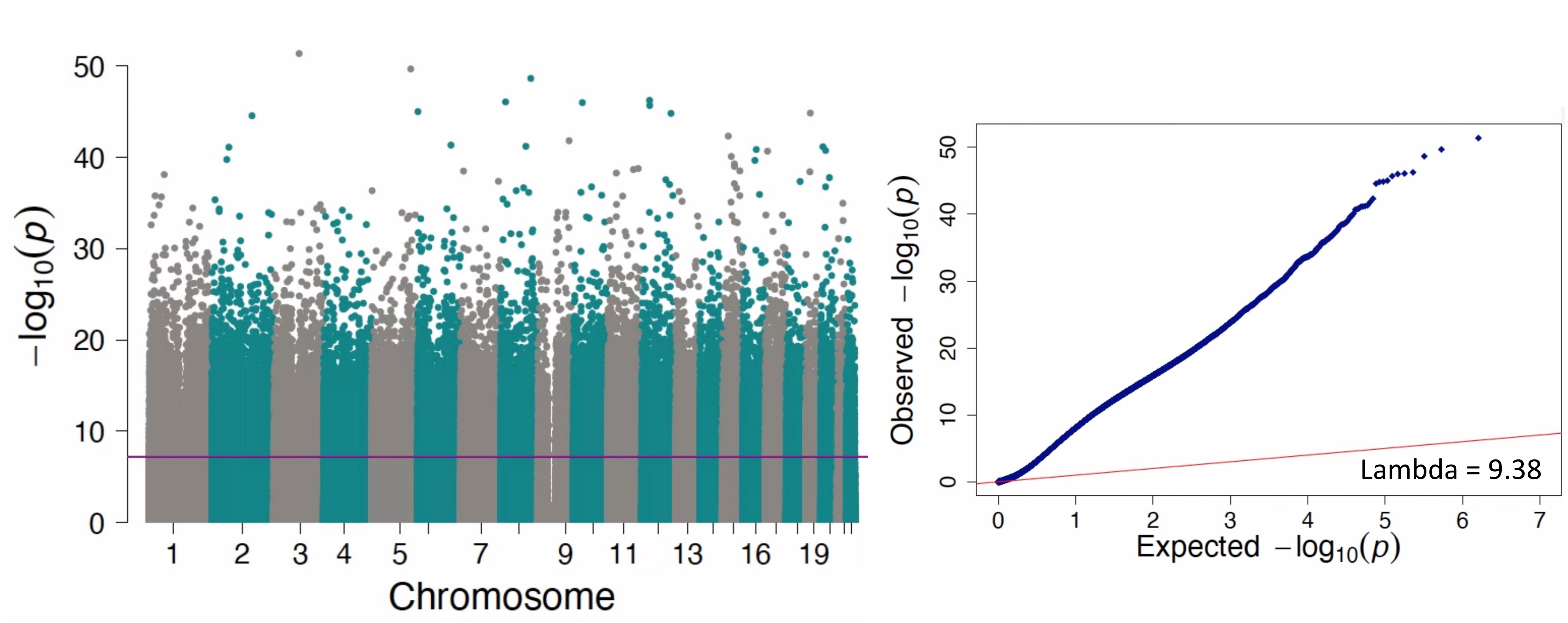


**Supplementary Figure 2: Epigenome-wide differential methylation analysis in CpGassoc identified DNAm signature associated with UC at diagnosis. The QQ plot representation shows huge inflation for the CpG sites associated to UC at diagnosis.**

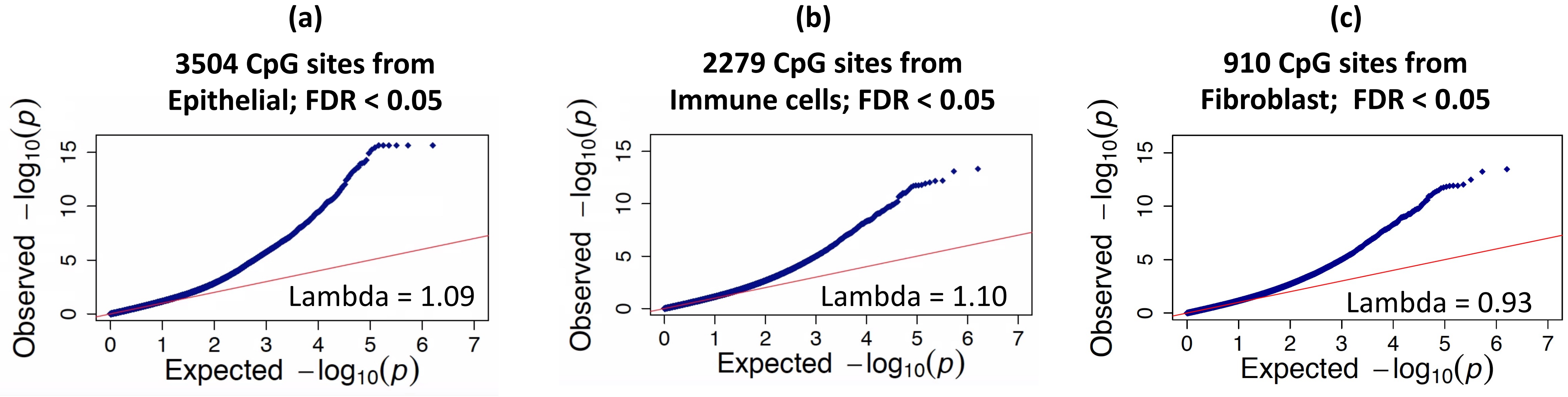


**Supplementary Figure 3: The QQ plot representation shows no inflation for the CpG sites associated to UC at baseline that are derived from (a) Epithelial and (b) immune cells, and (c) Fibroblast**

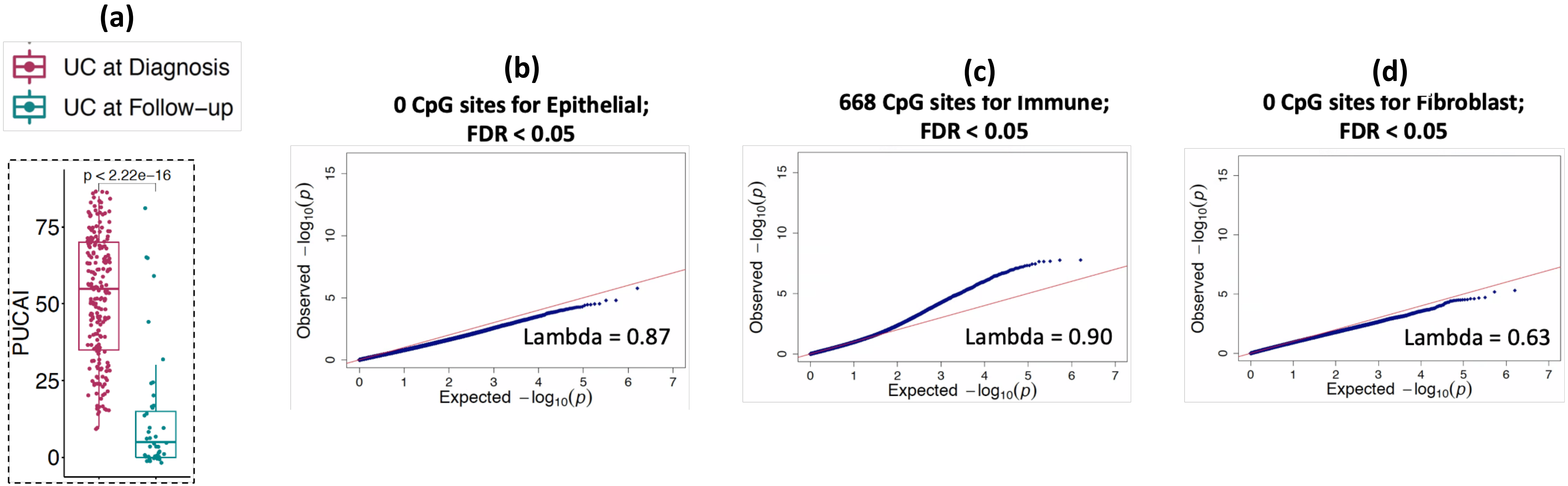


**Supplementary Figure 4: (a) Comparison between UC at diagnosis (n=211) and UC follow-up (n=73) for the pediatric ulcerative colitis activity index (PUCAI) showed a significant reduction in the follow-up UC. The P-value is calculated using the Wilcoxon test. (b-d) The QQ plot representation shows no inflation for the CpG sites associated to UC at disease course that are derived from both (b) Epithelial and (c) immune cells, and (d) Fibroblast**

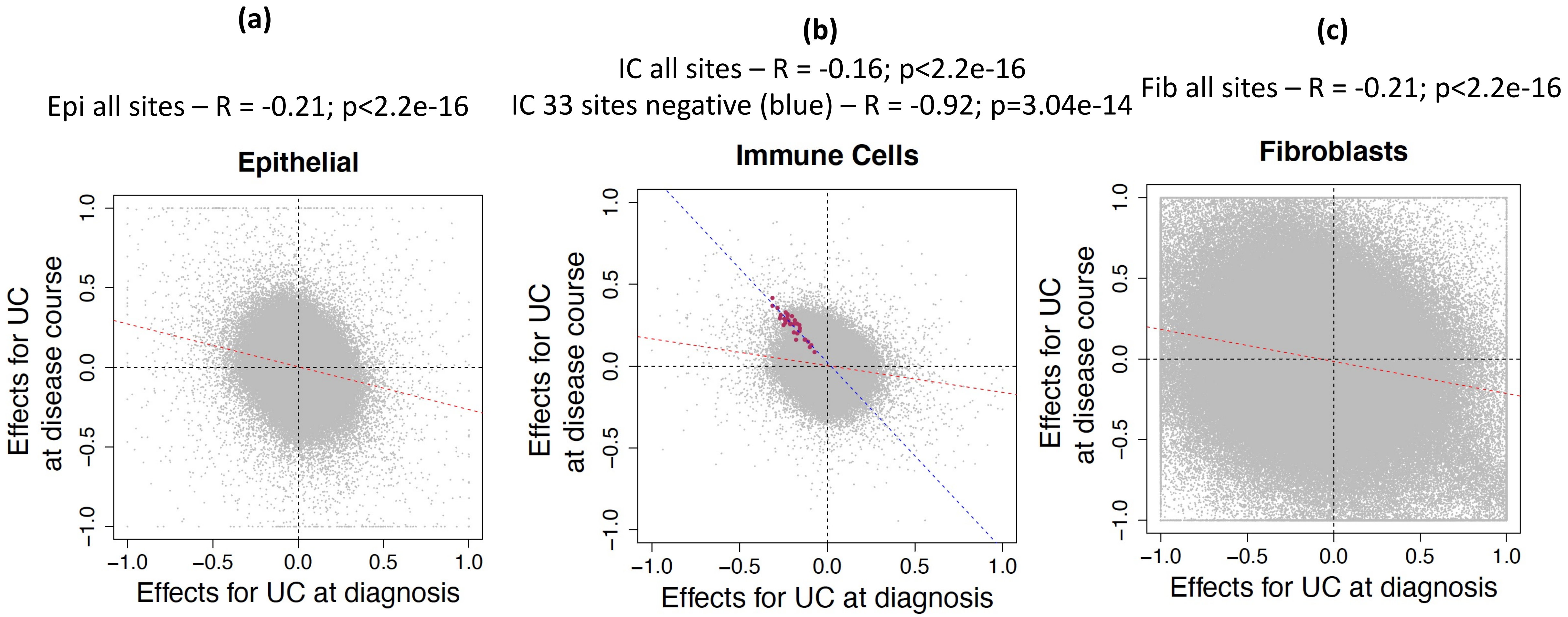


**Supplementary Figure 5: The effect sizes for entire 820K DNAm sites were compared between UC at diagnosis and UC at disease course (y-axis) for (a) Epithelial and (b) immune cells, and (c) Fibroblast. In B, maroon dots represents the immune signatures that are significant by FDR < 0.05 at both UC at diagnosis and UC at disease course.**

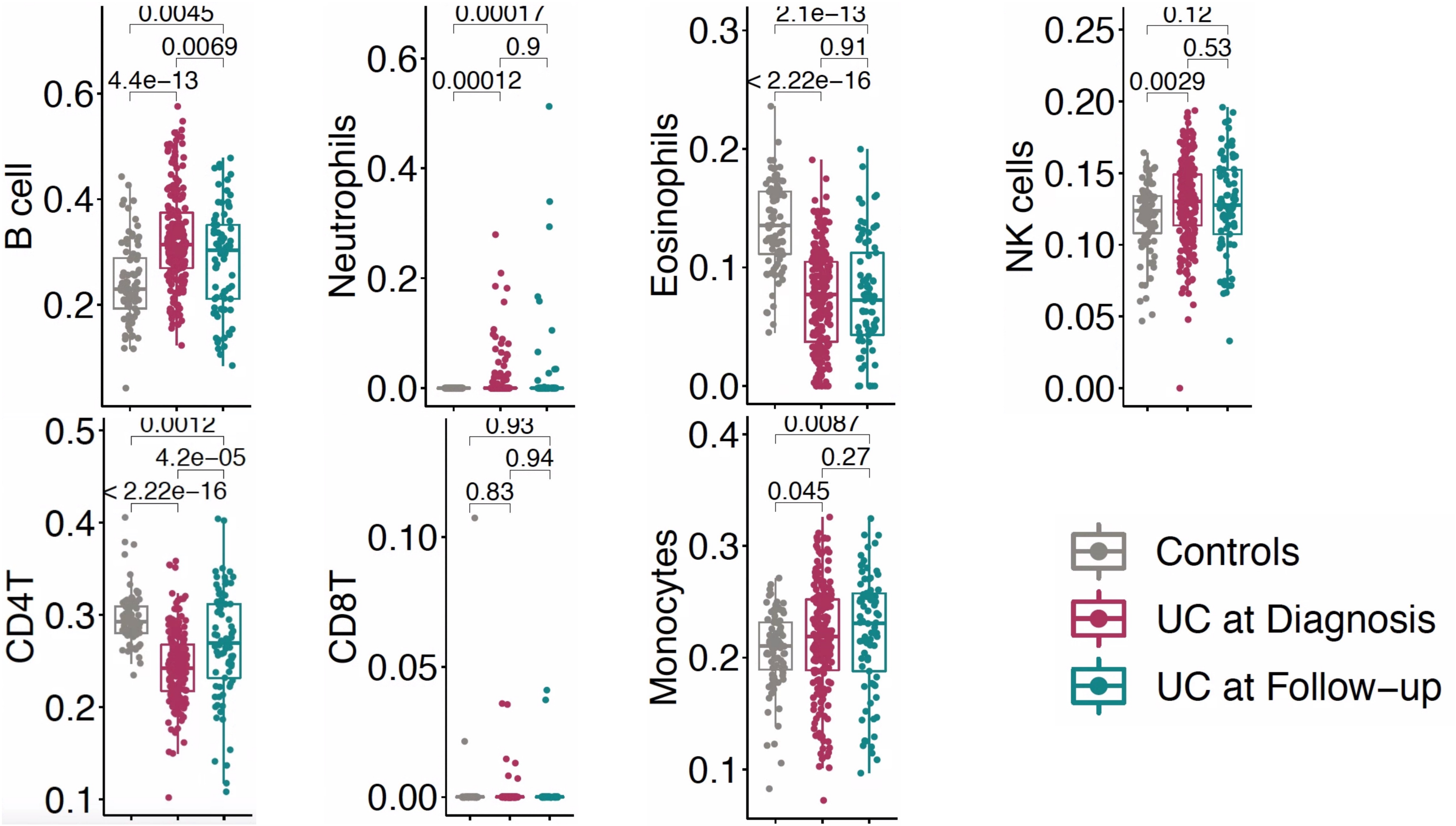


**Supplementary Figure 6: Overall estimated cell proportions for immune subtypes from rectal biopsy ~820K DNAm profile and they are compared among controls (n=85), UC at diagnosis (n=211) and UC at follow-up (n=73) are shown. X-axis shows the disease status and y-axis shows the estimated cell proportions from the Epidish CellDMC package. P-values are shown from the Wilcoxon test.**

**145 sites for Neutrophil FDR< 0.05**

**8 sites for B-cell FDR< 0.05**

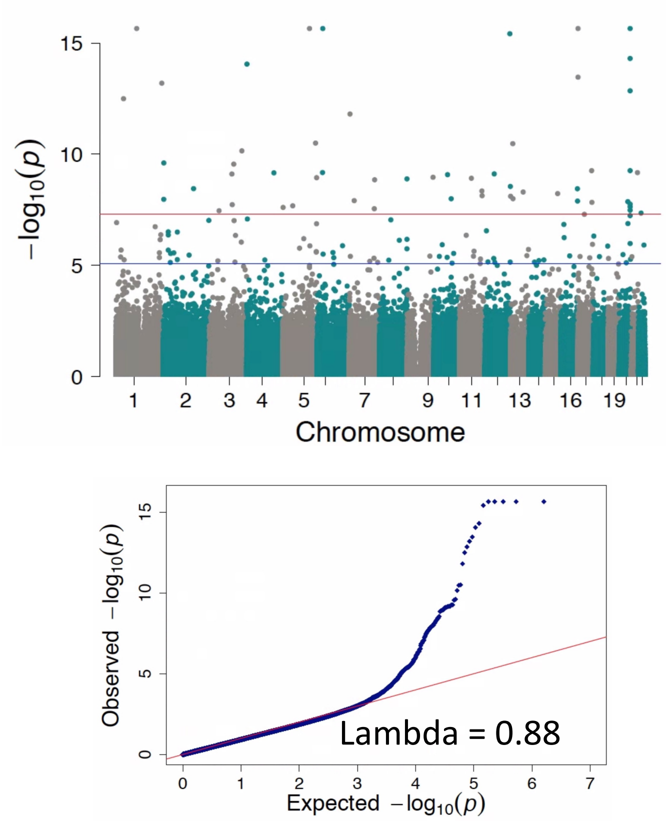


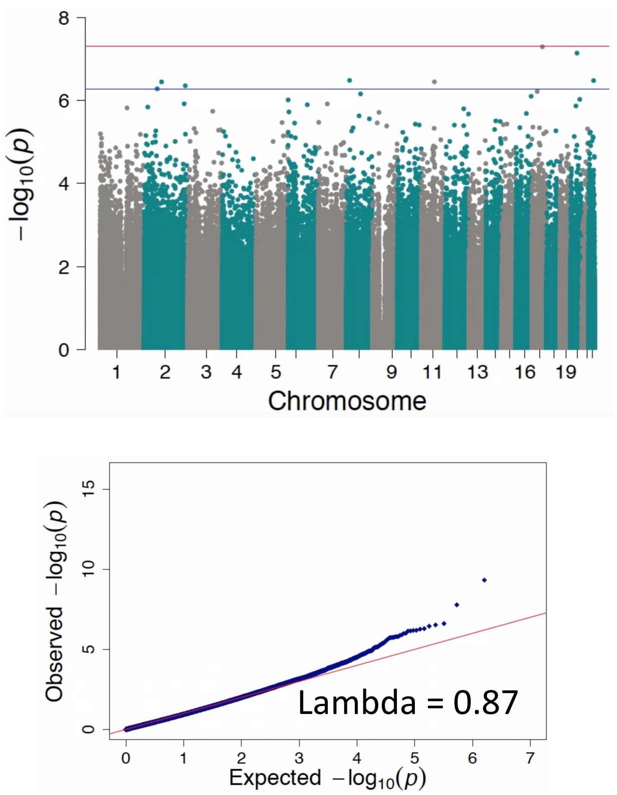


**Supplementary Figure 7: Epigenome-wide differential methylation analysis in CellDMC identified few signatures for two immune subtypes namely Neutrophils and B-cell between UC at diagnosis (n=73) and UC at follow-up (n=73).**

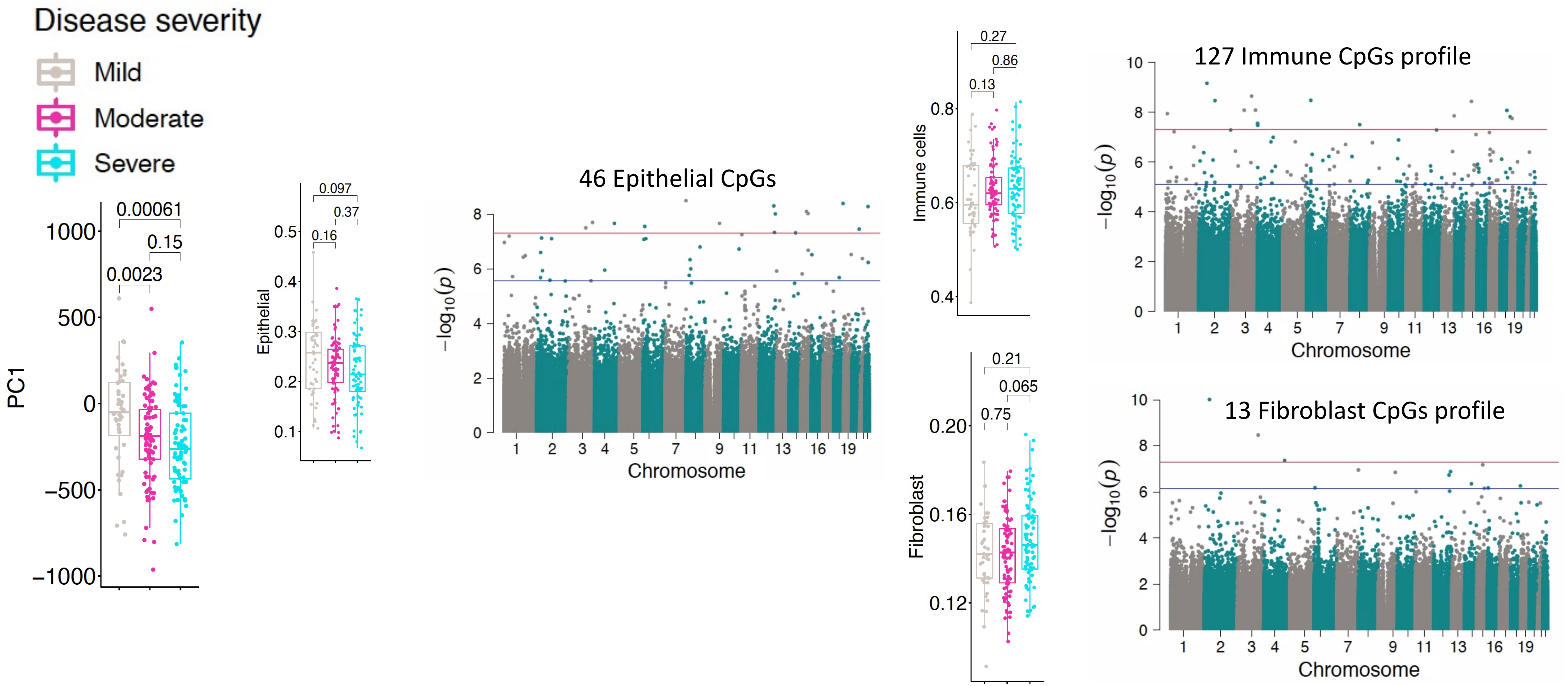


**Supplementary Figure 8: DNAm signatures associated with disease severity defined by PUCAI (A) The boxplots show the PC1 obtained from rectal biopsy DNAm profiles compared among mild UC (n=44), moderate UC (n=87) and severe UC (n=80) defined by PUCAI. Top panel shows the epithelial, immune, and fibroblast cell proportions obtained from entire 820K sites at genome-wide (B) Cell-specific EWAS analysis shows differentially methylated sites between for moderate and severe UC combined (n=167), when compared to mild UC. The results shown for all three major cell types from top to bottom for epithelial, immune and fibroblast, respectively. The blue line indicates the sites significant by FDR < 0.05 and the red line represents the sites significant by genome-wide p< 1e-08.**

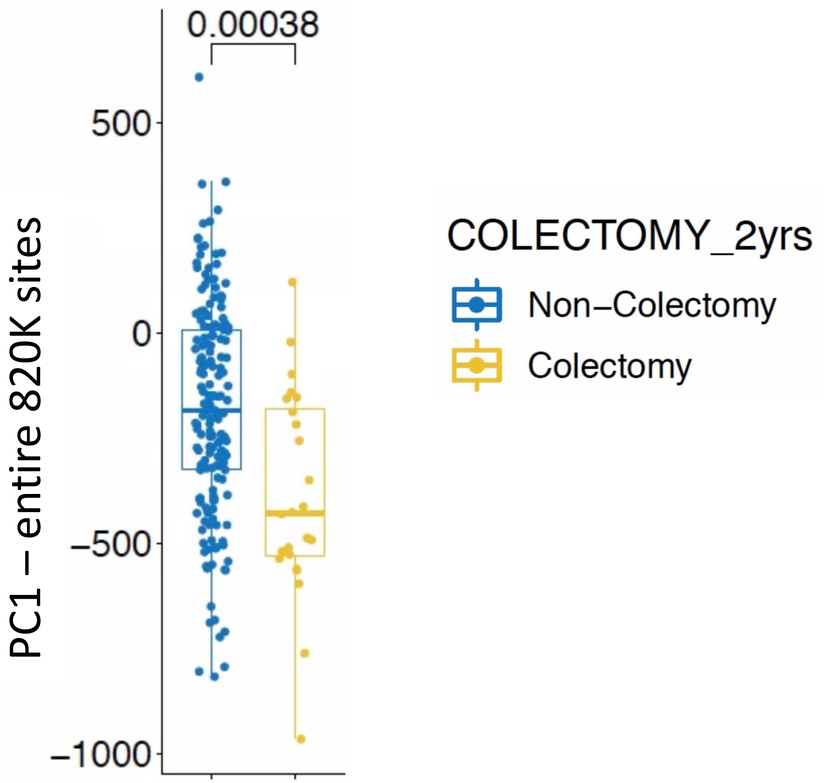


**Supplementary Figure 9: Boxplot comparing PC1 of entire 820K sites among colectomy groups at diagnosis. X-axis shows non-colectomy UC patients (n=175) and UC patients who undergone colectomy (n=24) at 2 yrs . Y-axis shows the PC1 values obtained from entire 820K array. The given p-value was calculated by Wilcoxon test.**

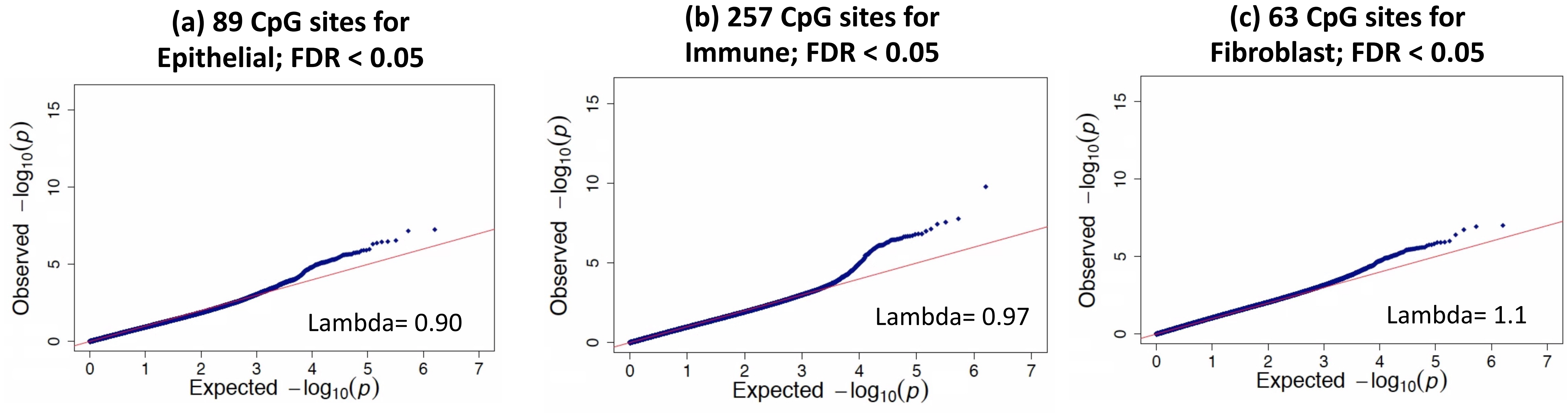


**Supplementary Figure 10: The QQ plot representation shows no inflation for the CpG sites associated to colectomy status in UC that are derived for (a) Epithelial and (b) immune cells, and (c) Fibroblast**

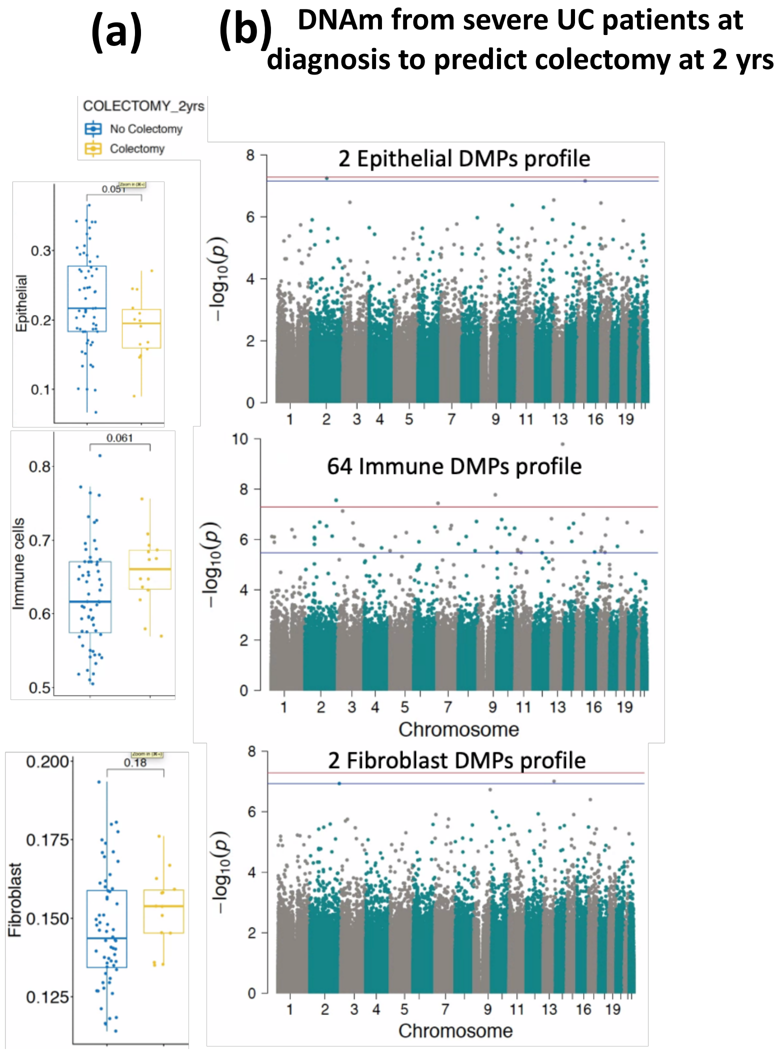


**Supplementary Figure 11: (a) The boxplots comparison of estimated cell proportions for epithelial, immune cells and fibroblasts obtained from rectal biopsies at diagnosis compared between colectomy from severe patients alone and non-colectomy UC severe patients. P-values are shown from the Wilcoxon test. (b) Cell-specific epigenome-wide DNAm analysis (EWAS) comparing colectomy from severe UC patients to non-colectomy severe UC patients. The blue line represents significant differential methylation with FDR < 0.05 and the red line represents Bonferroni-adjusted genome-wide significance (p< 1e-08).**

| | Samples | Cell types | | |
|---|---|---|---|---|
| | Training dataset (75%) | Epithelial | Immune | Fibroblast |
| UC vs Ctrl | 158 UC; 64 CTRL | 1010 | 80 | 39 |

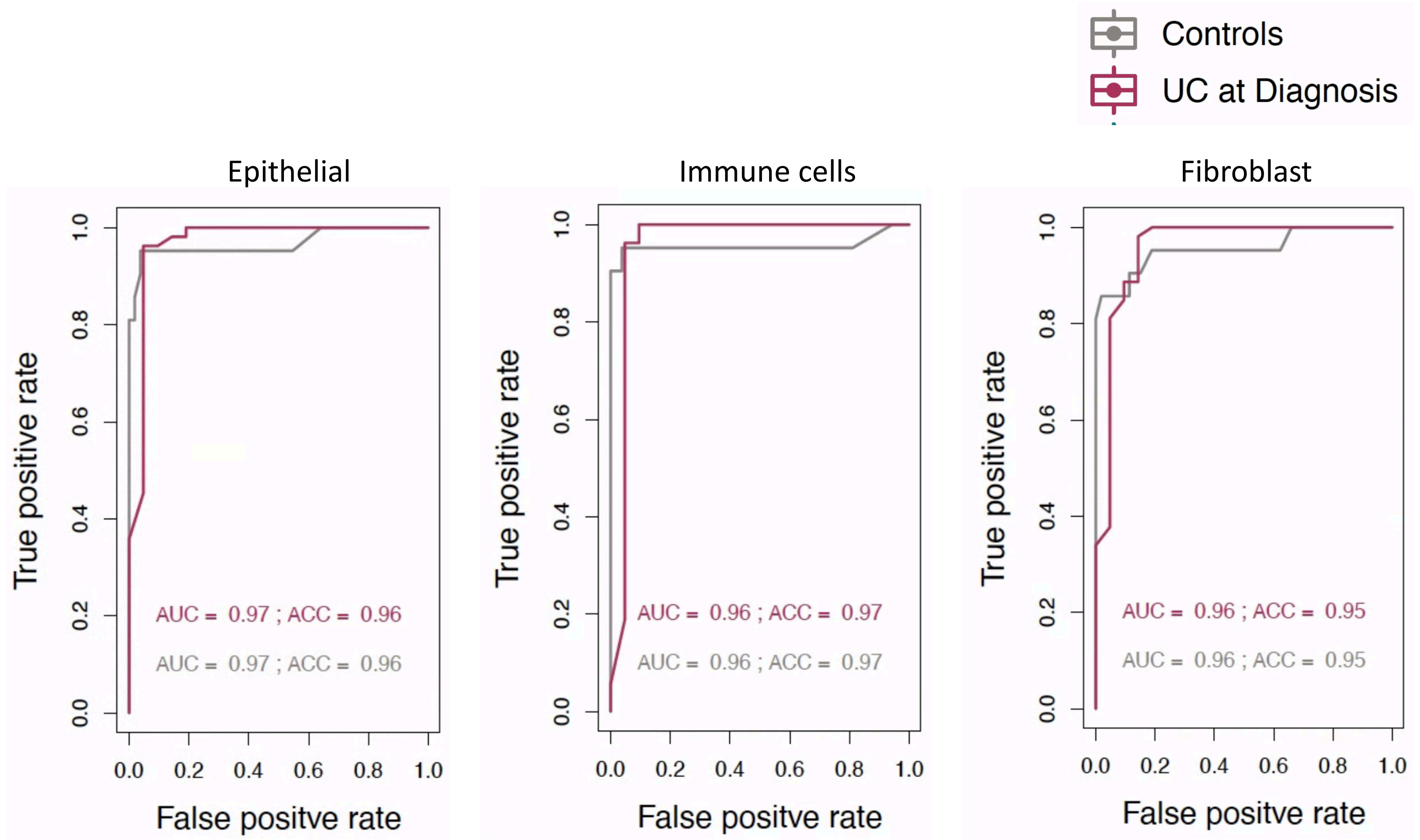


**Supplementary Figure 12: Clinical utility of rectal biopsy DNA methylation data for the prediction of case-control status. Random forest (RF) classification was performed on Epidish cell-specific EWAS signatures. RF model was trained on 75% training dataset and the same signatures were tested in the rest of 25% test dataset.**
